# Appetite and Energy Intake in Laboratory and Free-Living Conditions Remain Consistent Across Menstrual Cycle Phases when Using Precise Measurement Methods

**DOI:** 10.1101/2025.02.28.25323098

**Authors:** Miranda Smith, Maryam Aghayan, Jonathan Little, Jerilynn Prior, Tamara R Cohen, Zoë Soon, Hephzibah Bomide, Sarah Purcell

## Abstract

Self-reported dietary intake varies across menstrual cycle phases, but objective assessments of dietary intake together with appetite and resting metabolic rate (RMR) are limited. This study aimed to assess differences in appetite, dietary intake, and RMR during two hormonally-distinct menstrual cycle phases in laboratory and free-living settings.

Healthy premenopausal females with predictable normal-length menstrual cycles completed two study visits: one in the late-follicular and one in the mid-luteal phase. Menstrual cycle phases were identified using urinary luteinizing hormone surge and cycle days. Participants consumed a 2-day energy- and macronutrient-balanced run-in diet prior to each visit. RMR was measured with indirect calorimetry, followed by appetite ratings before and after a standardized breakfast, and completed a food cravings questionnaire. Appetite was also tracked for 2.5 days post-visit in a free-living environment. *Ad libitum* energy and macronutrient intake were measured using pre-weighed plus weighing of uneaten food at an in-laboratory lunch meal, as well as during the 2.5-day free-living period.

Eighteen participants were included (age: 21±4 years; body mass index: 21.2±1.5 kg/m^2^). There were no differences between in-laboratory *ad libitum* energy or macronutrient intakes, appetite, or food cravings between phases. RMR did not differ between phases, although the mid-luteal phase RMR trended toward higher (104±218 kcal/day higher*; P*=0.074). No main nor interaction effects for phase and time were observed for free-living dietary intake nor appetite ratings. Accurate measurements show no differences in appetite or energy intake between menstrual cycle phases, though RMR may be slightly elevated in the luteal phase.

## Introduction

Characterizing energy balance (i.e., dietary intake, energy expenditure) and its determinants is crucial for understanding the mechanisms of body weight regulation (1). Sex steroid hormones (i.e., estrogens and progesterone) may affect different aspects of energy balance parameters (2,3). This influence is particularly evident across the menstrual cycle, during which high estrogen and low progesterone occur in the late-follicular phase and moderate estrogen and high progesterone occur in the mid-luteal phase (4–6). These dynamic hormonal fluctuations may drive changes in appetite, dietary intake, and resting metabolic rate (RMR), contributing to variations in energy balance.

Previous research has found that hunger, dietary intake, and RMR tend be highest in the mid-luteal phase of the menstrual cycle (7–13). However, much of this research has relied on self-reported dietary intake, which is susceptible to bias and measurement error (14–16). Furthermore, many studies have used self-reported measures to identify menstrual cycle phases, which can misclassify phases and substantially impact the interpretation of energy balance in different hormonal states (8,10,17–26). Moreover, few studies have measured dietary intake together with energy expenditure, limiting the interpretation and generalizability of the findings within the broader framework of energy balance.

This study aimed to address these knowledge gaps by comparing appetite sensations, objectively-measured dietary intake, and RMR between late-follicular versus mid-luteal phases of the menstrual cycle in healthy premenopausal females. We hypothesized that hunger, prospective food consumption, dietary intake, and RMR would be higher, while satiety would be lower, in the mid-luteal phase compared to the late-follicular phase.

## Methods

### Participants

Participants were recruited from the greater Kelowna community from May 2022 – September 2024. Healthy, premenopausal females aged 18-35 years with normal body weight (18.5-24.9kg/m^2^) were eligible. Other major inclusion criteria included: nulliparity; regular normal-length and ovulatory menstrual cycles, as indicated by self-reported menstrual cycle length between 21-38 days for the past 3 cycles and ovulation identified via urinary luteinizing hormone (LH); being sedentary or recreationally active, defined as < 300 minutes of moderate or vigorous intensity exercise per week; not currently pregnant or lactating or planning to become pregnant or lactate in the next 12 weeks; and ability to fast for 12 hours prior to each study visit. Major exclusion criteria included: presence of a major chronic disease (e.g., cardiovascular diseases, diabetes, cancer, thyroid disease, etc.); use of oral contraceptive pills or any hormone-related medication; progestin-releasing intrauterine device; or medications that may affect appetite, energy balance, or sleep in the past six months. Individuals who used tobacco or nicotine, worked night shifts, had a history of extensive weight loss or weight loss surgery, or had food intolerances or allergies that could not be accommodated were also excluded. Other exclusion criteria included current or past history of eating disorders including anorexia nervosa, bulimia, binge eating disorder (self-report or score >20 on the Eating Attitudes Test – 26 questionnaire (27)), current severe depression or history of severe depression within the previous year (self-report or 30 score > 30 on Beck Depression Inventory (28) or alcohol or drug abuse (score >1 or >2 with prior principal investigator approval on the cut-annoyed-guilty-eye-opener (29)). The project was approved by the University of British Columbia Clinical Research Ethics Board (ID: #H22-00874) and all participants provided signed consent prior to the study.

### Study design

This study consisted of a screening and baseline visit and two study visits – one in the late-follicular phase and one in the mid-luteal phase of the menstrual cycle. Participants were provided with a two-day energy-balanced diet before each study visit day. Appetite sensations, objective (weighed) dietary intake, and RMR were measured during each of these study visits as described below. Appetite sensations and objective dietary intake were also measured for 2.5 days following each study visit.

### Screening and baseline visit

Individuals who were eligible and provided written informed consent completed a screening and baseline visit to familiarize participants with the physical space in which they would complete the study visits, measure body composition, assess menstrual cycle history, and review instructions for measuring ovulation. Height and weight were measured using a stadiometer and digital scale, respectively. Percent body fat was measured using dual-energy x-ray absorptiometer (Hologic, Horizon DXA system, Auto Whole-Body Fan Beam, version 13.4.2., Bedford, MA, USA) to characterize the population.

Each participant provided their menstrual cycle history using a study-specific form to assess menstrual cycle consistency for the past three months and to predict future ovulation and menstrual cycle phases. At the end of the screening and baseline visit, participants were given LH urine test strips with disposable collection cups to prospectively identify ovulation. Using their previous menstrual cycle history, participants were asked to use the LH tests each day during a 7-day period in which ovulation was likely to occur. A positive ovulation test was used to ‘anchor’ study visits, with the late-follicular phase study visit scheduled 1-5 days before the next predicted ovulation date and the mid-luteal phase study scheduled 6-10 days after the estimated ovulation date. A minimum of one positive urinary LH test was required prior to completing any study visit. Participants were asked to track their LH throughout the entire duration they were enrolled throughout the study.

### Experimental sessions

#### Run-in energy balanced diet

Because dietary intake in previous days may impact appetite and dietary intake on subsequent days (30), participants were provided with food to support a run-in energy-balanced diet in free-living settings two days before each study visit. The energy content of the run-in diets was calculated using the Dietary Reference Intakes (31) with the physical activity coefficient selected based on each participant’s self-reported activity level (collected at the screening and baseline visit). The macronutrient content of the run-in diet was 50% carbohydrates, 30% fat, and 20% protein. During this period, participants were asked to abstain from consuming any foods not provided to them, but were permitted to consume beverages (e.g., tea, coffee, diet soda) if it did not contain energy. If participants did consume food or calorie-containing beverages not provided to them, they were instructed to send detailed information about the food (e.g., the nutrition label, photos, description) via email and return the packaging to the study team.

Participants were given the same energy- and macronutrient-adjusted meals and snacks to consume before each study visit. Four meals were prepared by a local third-party company in Kelowna (Meal Prep 4 U), which were supplemented with a breakfast meal (i.e., a combination of peanut butter and jelly bagels, granola with yogurt, toast with butter or jam, and egg bites) and various snacks (e.g., granola bars, apples, bananas, trail mix, hummus, yogurt, juice boxes) for each day, based on participant preferences and estimated energy requirements.

### Study visits

Before each study visit, participants were asked to refrain from vigorous exercise for 48 hours, avoid alcohol for 24 hours, and avoid calorie or caffeine-containing food and beverages for 12 hours. Each study visit was scheduled to begin between 7:30-9:30 am, with similar start times (within one hour) for each study visit.

RMR was measured using an indirect calorimeter with a face mask (Parvomedics TrueOne 2400, Murray, Utah). Gas analyzers and the flow meter were calibrated prior to each test according to manufacturer’s instructions. Before each test, participants lay supine in a quiet, thermoneutral (20-24°C) room for 20-25 minutes. Respiratory gas exchange was measured for 20-25 minutes. The last 15 minutes of data were averaged after inspection and exclusion of data that did not meet quality control targets (i.e., minute-by-minute coefficient of variation <10% for volume of O_2_ and CO_2_ and RMR).

Following RMR testing, participants rated sensations of hunger, satiety, and prospective food consumption (PFC) (32) using sliding VAS, which were covertly scored on a scale of 0-100 on a computer or iPad using Research Electronic Data Capture (REDCap). These ratings were completed again 30, 60, 90, 120, 150, and 180 minutes after the breakfast meal. Between VAS and questionnaires, participants could complete light work on their computer, read, work, or watch television and were asked to do similar activities at each study visit. Area under the curve was calculated to describe differences in appetite sensations between menstrual cycle phases using the trapezoidal method (33). After the 120-minute VAS assessment, participants completed the 15-item Food Cravings Questionnaire – State (34). Higher scores indicated more intense food cravings in that moment.

After the fasted VAS, participants were given a standardized breakfast meal consisting of 25% of the individual’s total daily estimated energy requirements (31) and the same macronutrient ratio as the run-in diet (50% carbohydrates, 30% fat, and 20% protein). Each breakfast consisted of a combination of toast, butter, egg quiche, fruit, yogurt and/or juice, depending on participant preference and energy and macronutrient requirements. Participants consumed this meal with no distractions to avoid influence from external stimuli. Participants were permitted to drink water *ad libitum* during and after the breakfast meal. At the end of the 180-minute VAS, participants were provided with an in-laboratory lunch meal in which each food was weighed before and after consumption to objectively measure *ad libitum* dietary intake. This meal provided approximately 1800 kcal, of which approximately 60% were from carbohydrates, 30% were from fat, and 10% were from protein. Items included spaghetti, meatballs (or non-meat alternative), marinara sauce, a green vegetable with butter (e.g., broccoli) or green leafy vegetables, Italian dressing, ranch dressing, a medium apple, a packet of cookies, regular soda, and diet soda. For this meal, participants were asked to consume as much or as little as they would like until they were comfortably full and could request more of any item. This meal was consumed in a quiet room with no external distractions.

At the completion of each study visit, participants began their free-living period, in which they were provided pre-weighed food in excess of their estimated energy requirements for the remainder of the study visit day and for two subsequent days. The provided foods consisted of 4-6 pre-prepared meals with known energy and macronutrient content from a local company, quick breakfast and alternative meal options (e.g., bread and frozen egg quiches, instant macaroni and cheese) and several fresh and packaged foods that could serve as snacks or meal supplements (e.g., fresh and canned fruit, granola bars, chips, assorted candies, oatmeal packets, egg bites, brown and white rice, mac and cheese). While the content of the diet varied slightly based on participant preferences, the total energy provided was approximately 10,500 kcal (∼4,200 kcal/day). Similar to the in-laboratory lunch meal, participants were instructed to consume as much or as little of the foods and beverages provided as they wanted and exclusively consume foods provided by the laboratory. Participants were provided with the same foods for each free-living *ad libitum* period. To confirm *ad libitum* dietary intake, participants were instructed to take before and after photographs of all food and beverages they consumed using their phone. Photos were uploaded directly to REDCap using a personal link, which recorded the time in which foods were consumed. At the end of the 2.5-day period, participants were asked to bring back all food and wrappers/containers so all items could be weighed by the research team. Dietary energy and macronutrient intake were assessed using Food Processor Nutrition Analysis Software (version 11.1; ESHA Research, Salem, OR), with entries confirmed by a research assistant. Daily free-living energy intake was expressed as kcal/day and adjusted for RMR (energy intake / RMR, kcal/day) To explore free-living appetite, participants also provided VAS of hunger, satiety, and PFC in REDCap before and after each meal or snack, which were averaged across days.

### Statistical analyses

The sample size for this study was determined based on anticipated differences in-laboratory *ad libitum* dietary intake between the late-follicular and mid-luteal phases of the menstrual cycle (primary objective). We anticipated an effect size of 0.800, consistent with previous research using similar methods that detected differences in energy intake at a single meal between menstrual cycle phases that would be relevant for future nutrition strategies or care (i.e., 141 ± 176 kcal at a single meal (8)). A sample size of n = 14 would provide 80% power to detect mean differences in energy intake between phases using a two-tailed paired t-test with a significance level (α) of .05.

Statistical analyses were conducted using SPSS (IBM SPSS Statistics, version 29.0.00.0, Chicago, IL, USA). Distribution of data was assessed using the Shapiro-Wilk test of normality. Linear mixed effect models with unstructured covariance compared outcomes between menstrual cycle phases while maximizing statistical power in the presence of missing data. Models included participants as random effects and menstrual cycle phase (late-follicular or mid-luteal) as a fixed effect; in the case of VAS and free-living appetite and dietary intake data, time (fasting, 30, 60, 90, 120, 150, and 180 minutes for VAS; days 1, 2, and 3 for free-living data) and their interaction terms were also included as fixed effects with an unstructured covariance structure. Models with free-living appetite ratings included free-living energy intake as a covariate. In cases where model residuals were not normally distributed, data were log-transformed and re-analyzed, but absolute mean values were presented for ease of translation. Data are presented as mean ± SD or mean ± SE unless indicated otherwise.

## Results

### Participant characteristics

Baseline characteristics of study participants are shown in **Table 1**. Thirty-one females were enrolled, although 13 were excluded before the first study visit due to scheduling conflicts (n = 6), exclusionary survey score (n = 3), dietary restrictions (n = 2), and non-response (n = 2). Of the remaining 18 participants, 2 completed one visit. Four individuals had missing data for in-laboratory *ad libitum* dietary intake, 7 had missing data for ad libitum dietary across a 2.5-day free-living period and 6 had missing RMR data in one of the visits. There was complete data from 14 participants for our primary outcome (in-laboratory *ad libitum* dietary intake). Data from all 18 participants were included in analyses. All 18 participants had a positive ovulation test (i.e., confirmed by a LH urine test) prior to their first visit. Eight participants had their first visit in the late-follicular phase.

**Table 1.** Participant characteristics (n = 18).

| Characteristic | Mean ± SD or n (%) |
| --- | --- |
| Age (years) | 21 [19-24]* |
| Race |  |
| Black | 1 (6%) |
| White | 13 (72%) |
| East Asian | 2 (11%) |
| Middle Eastern | 2 (11%) |
| BMI (kg/m<sup>2</sup>) | 21.2 ± 1.5 |
| Percent body fat | 25.0 ± 4.5 |
| Mean cycle length (days) | 30.7 ± 3.0 |
| Number of confirmed ovulation tests |  |
| 1 | 5 (28%) |
| 2 | 10 (56%) |
| 3 | 2 (11%) |
| 4 | 1 (6%) |

\*\*Age (years) reported in median [interquartile range]. All other data are reported as mean ± SD or number (%). BMI: body mass index.

### Appetite sensations

VAS ratings of in-laboratory hunger, satiety, and PFC are shown in **Figure 1** and **Supplementary Table 1**. As expected, there was a main effect of time on all VAS (all *P*<0.05). There were no main effects of phase or phase x time interactions among the VAS, nor were there effects of phase on AUC values of appetite or food cravings score (**Supplementary Table 2**).

**Figure 1.**
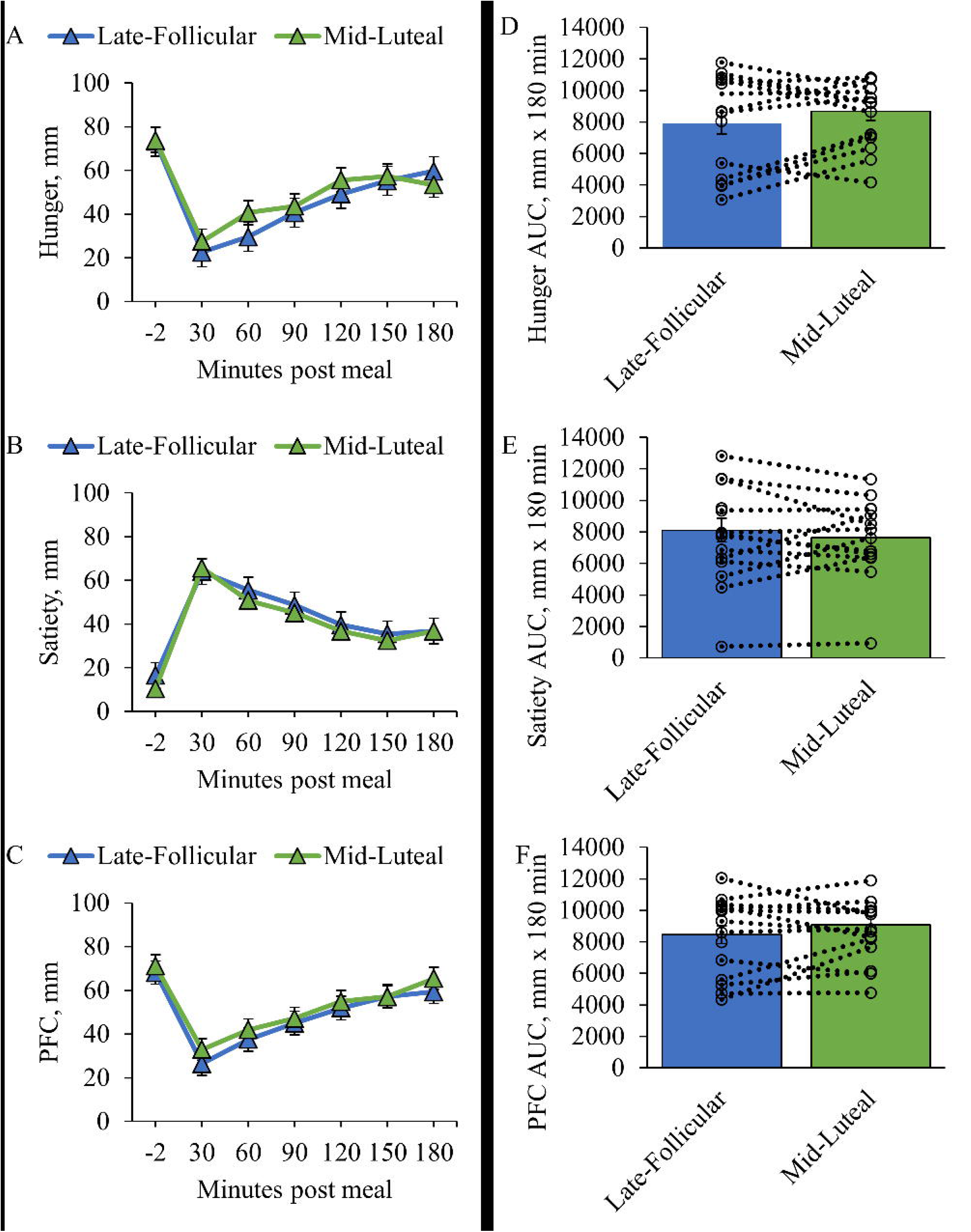
Time course change of hunger (A), satiety (C), and prospective food consumption (PFC; E) after a standardized meal. Area under the curve (AUC) for hunger (B), satiety (D), and PFC (F) are also included. Values are mean ± SE. N = 17 late-follicular, n = 17 mid-luteal.

There were also no differences in free-living ratings of hunger, satiety, or PFC (**Supplementary Table 3**).

### Dietary intake and resting metabolic rate

There were no differences in-laboratory *ad libitum* energy intake (**Figure 2**) or macronutrient intake (**Supplementary Table 2**) between menstrual cycle phases. There were also no main (phase, time) or interaction (phase x time) effects for free-living dietary intake, **Supplementary Table 2** and **Supplementary Table 4**. There were no differences in RMR between phases, although there was a trend towards greater RMR in the luteal phase (104 ± 218 kcal higher in the luteal phase, *P*=0.074, **Supplementary Table 2**).

**Figure 2.**
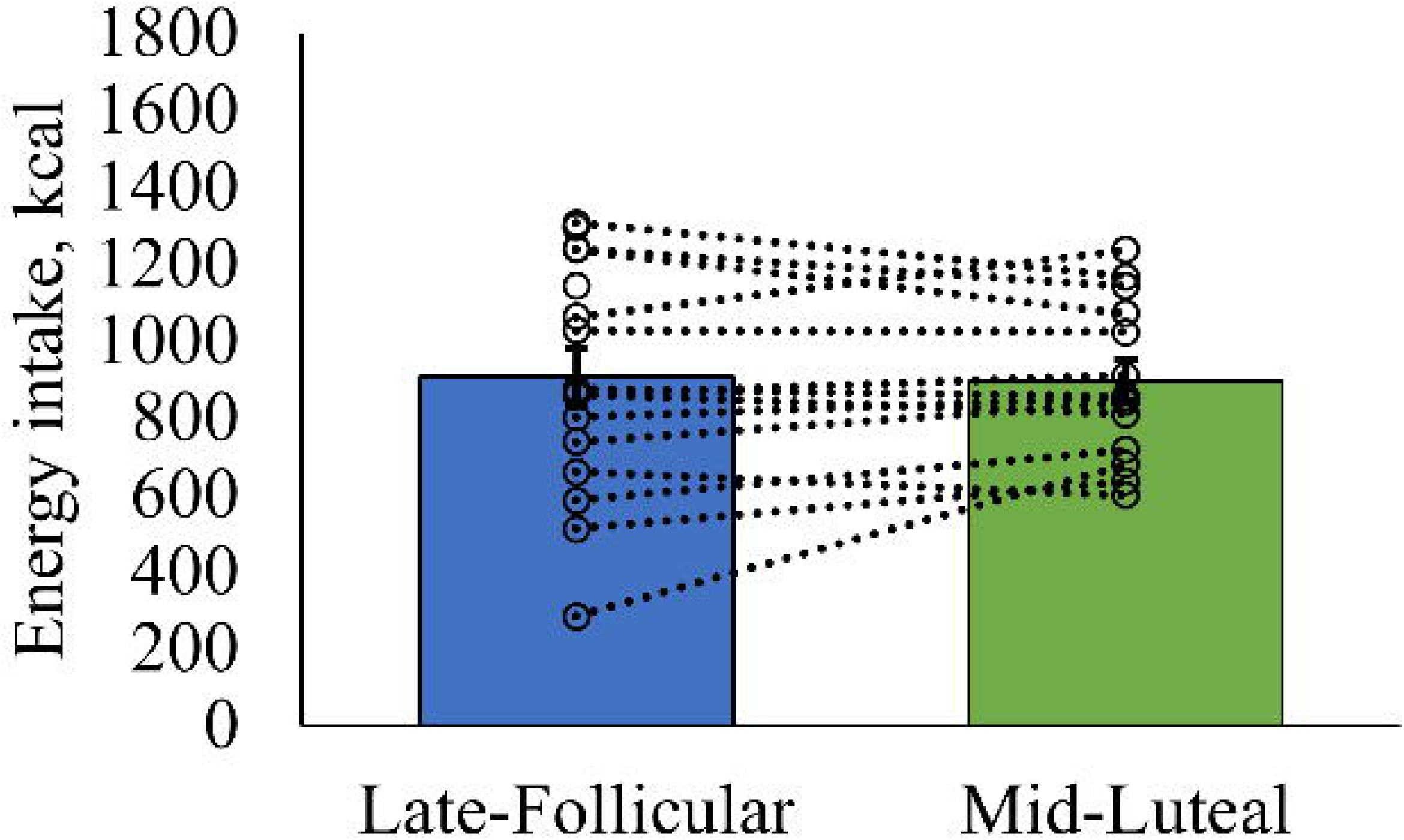
Mean energy intake (kcal) at a single meal between the late-follicular and mid-luteal phases of the menstrual cycle. Values are mean ± SE. N = 16 late-follicular, n = 14 mid-luteal.

## Discussion

This study challenges the prevailing notion that appetite and energy intake consistently increase during the luteal phase of the menstrual cycle. Despite common claims in previous research that suggest hormonal fluctuations lead to heightened orexigenic appetite sensations and greater energy intake in the luteal phase (8,10,12,17,19,22–25,35–37), our findings show no significant differences in energy intake or appetite sensations, whether measured in controlled laboratory conditions or real-world settings. This suggests that the menstrual cycle phase may not exert as robust an effect on energy intake as previously suggested, particularly when both menstrual phase and energy intake are measured with high accuracy.

Our findings suggest that differences in dietary intake reported in previous studies ma not be solely due to biological differences, but also differences in study design, particularly in the methods used to assess dietary intake and menstrual cycles. A major factor influencing discrepancies across studies is the reliance on self-reported dietary intake, which is prone to underreporting and bias (15). Self-reported energy intake has been estimated to be up to 522 kcal/day lower in the follicular phase compared to the luteal phase (22). Interestingly, studies using self-reported methods, such as 24-hour dietary recalls and food diaries, often report higher energy intake during the luteal phase (17,19,20,22,24,25,35–37), while studies utilizing weighed food intake methods report smaller (87-207 kcal/day) or no differences (10,23). While speculative, it is possible that some of this discrepancy may be due to societal perceptions of dietary intake across the menstrual cycle and media messages suggesting that it is normal for females to consume more before menstruation because of pre-menstrual syndrome may contribute to self-reported energy intake inaccuracies.

Another important consideration for interpretation of our data in the context of the wider literature is menstrual phase classification. Many prior studies have relied primarily on self-reported onset of menses to determine menstrual cycle phase (8,10,17–26), which can be inaccurate due to variability in cycle length, ovulation, and subclinical ovulatory disturbances (38,39). This is particularly relevant because relying on a fixed 28-day cycle or averaging previous cycles may lead to significant misclassification of menstrual cycle phase. Even small variations in cycle length can lead to substantial shifts in hormone levels, potentially altering the conclusions drawn from the data (40). For example, in a study of 39 healthy premenopausal women, excluding participants with misclassified cycle phases (based on self-report vs. retrospective serum hormone analysis) revealed higher energy and protein intake during the luteal phase (41). This study showed that 44% of cycles could be misclassified when relying on cycle history without more objective techniques, and that including such misclassified cycles may alter conclusions about phase-related differences in energy intake. Studies that objectively confirm menstrual phase through hormonal verification, as done in this study, are likely to yield more precise comparisons. Additionally, failure to account for previous day’s dietary intake and providing the same breakfast meal to all participants before postprandial appetite and energy intake assessments can introduce errors, as prior dietary intake (30) and individual differences in energy requirements are key factors influencing subsequent intake (42).

No significant differences in appetite sensations were observed in either a controlled laboratory or free-living environment, as assessed by VAS. Despite the trends generally aligning with expectations—namely, increased hunger in the luteal phase (8) — our findings are consistent with other research showing no significant difference in appetite VAS between phases (9,11). However, it is important to consider that VAS may not be sensitive enough to capture the full complexity and depth of appetite regulation (43). Furthermore, appetite sensations may not directly relate to self-reported energy intake (44). In free-living conditions, which are inherently challenging to quantify, no phase-specific differences were detected, although the methodology used to assess free-living appetite in this study has not been formally validated.

Previous studies have reported differences in various components of appetite related to food cravings across the menstrual cycle. For example, some evidence suggests that females experience a stronger preference for high-fat and high-sugar foods during the late-luteal phase (45) and a greater desire for foods rich in sugar, salt, and fat during the premenstrual period (26). Increased explicit wanting for high-fat foods (46), higher consumption of sweet foods (47), and greater intake of sugar-containing beverages (21) have also been reported in the luteal phase, potentially driven by hormonal fluctuations, such as elevated progesterone levels (48). Despite these findings, the current study did not observe significant differences in food cravings between phases, although the trends were in the expected direction. This may be due to the limitations of the Food Craving Questionnaire – State, which only partially measures aspects of food cravings, particularly those linked to hedonic or emotional responses, and does not comprehensively account for all dimensions of appetite. Additionally, the insufficient statistical power for this specific portion of the study may have limited the ability to detect significant differences.

We observed a non-significant increase in RMR during the luteal phase that was approximately 100 kcal/day higher than in the follicular phase. Previous studies have reported mixed findings, showing differences ranging from no effect to approximately 50–100 kcal/day greater in the luteal phase (7,20,49). This increase may be attributed to the thermogenic effects of progesterone, which is greatest in the mid-luteal phase(50). For context, energy intake in the current was approximately 200 kcal/day higher in the luteal phase (although not significant), exceeding the observed increase in RMR. Persistent discrepancies in energy balance over prolonged periods could potentially lead to weight gain, though it remains unclear whether fluctuations in endogenous sex hormones directly influence long-term weight regulation, or if the increased energy balance observed in the luteal phase is counterbalanced by compensatory mechanisms during the follicular phase.

Strengths of this study include the use of objective and accurate methods to identify menstrual cycle phases and assess dietary intake in both controlled laboratory and free-living settings, alongside measurements of appetite and RMR—representing a novel approach with rigorous methodology. However, some limitations should be noted. The study was powered only to detect differences in in-laboratory energy intake, although many previous studies have used comparable or smaller sample sizes. Additionally, hormonal markers of appetite, which may vary across menstrual phases, were not measured. During the 2.5-day period, participants were provided with meals and snacks exceeding their caloric needs to minimize boredom, but some evidence suggests that providing large portions of food may lead to spontaneous overeating. Moreover, the study focused solely on healthy, young females with a normal BMI, limiting its generalizability to other populations.

In conclusion, this study challenges the assumption that appetite and energy intake consistently increase during the luteal phase of the menstrual cycle. Despite general trends aligning with previous research—such as increased hunger, energy intake, and RMR during the luteal phase—no significant differences were observed in either laboratory or free-living settings. These findings suggest that menstrual phase may not have as strong an effect on energy intake as previously thought, particularly when both cycle phase and dietary intake are assessed accurately. These results underscore the importance of refining measurement techniques and highlight the need for further research to explore the complex relationship between hormonal fluctuations and appetite regulation.

## Data Availability

All data produced in the present study are available upon reasonable request to the authors.

## Acknowledgements

We would like to sincerely thank Dr. Ali McManus and Dr. Neil Eves at the University of British Columbia Okanagan for their generous support in providing access to their equipment for the study visits.

## Author contributions

Sarah Purcell, Jonathan Little, and Jerilynn Prior designed the study. Miranda Smith and Maryam Aghayan contributed to the data collection. Tamara Cohen and Zoë Soon provided critical input for this study’s methods. Hephzibah Bomide contributed to the data analysis. Miranda Smith was responsible for most of the data collection and analysis and wrote the first draft of the manuscript. All authors edited subsequent drafts and approved the final article.

