## Supplementary for "Appetite and Energy Intake in Laboratory and Free-Living Conditions Remain Consistent Across Menstrual Cycle Phases when Using Precise Measurement Methods"

### Supplementary materials

**Supplementary Table 1. Descriptive data and analyses examining main and interaction effects of time and phase on repeated measures of appetite collected in the laboratory**

| | Time | | | Phase | | | Time x Phase | | |
|---|---|---|---|---|---|---|---|---|---|
| | ***df*** | ***F*** | ***p*** | ***df*** | ***F*** | ***p*** | ***df*** | ***F*** | ***p*** |
| **Hunger, mm x 180 min** | 6, 15.85 | 54.359 | <0.001 | 1, 16.49 | 1.784 | 0.200 | 6, 15.29 | 1.483 | 0.248 |
| **Satiety, mm x 180 min** | 6, 17.03 | 44.018 | <0.001 | 1, 16.81 | 2.066 | 0.169 | 6, 16.74 | 0.432 | 0.847 |
| **PFC, mm x 180 min** | 6, 13.84 | 57.740 | <0.001 | 1, 12.66 | 1.425 | 0.254 | 6, 13.52 | 2.237 | 0.103 |

Time is in increments of 30 minutes (for 180 minutes). PFC: prospective food consumption.

**Supplementary Table 2. Descriptive data and analyses examining main effect of phase on dietary intake at an in-laboratory lunch meal, appetite area under the curve (AUC), and resting metabolic rate**

| | **Late-follicular Mean ± SD** | **Mid-luteal Mean ± SD** | ***df*** | ***F*** | ***p*** |
|---|---|---|---|---|---|
| ***Dietary intake*** | | | | | |
| In-laboratory energy intake, kcal | 901 ± 294 | 895 ± 205 | 1, 15.31 | 0.256 | 0.620 |
| In-laboratory carbohydrate intake, g | 121 ± 41 | 122 ± 28 | 1, 16.42 | 0.113 | 0.741 |
| In-laboratory fat intake, g | 31 ± 13 | 30 ± 9 | 1, 13.06 | 0.328 | 0.576 |
| In-laboratory protein intake, g | 36 ± 12 | 36 ± 9 | 1, 14.23 | 1.034 | 0.326 |
| Free-living energy intake, kcal/day | 1994 ± 430 | 2198 ± 401 | 1, 8.46 | 1.212 | 0.301 |
| Free-living energy intake, RMR-adjusted, kcal/day | 1.68 ± 0.36 | 1.67 ± 0.42 | 1, 6.90 | 0.031 | 0.865 |
| Free-living carbohydrate intake, g/day | 237 ± 54 | 262 ± 44 | 1, 10.31 | 2.046 | 0.182 |
| Free-living fat intake, g/day | 78 ± 19 | 87 ± 21 | 1, 10.86 | 0.939 | 0.354 |
| Free-living protein intake, g/day | 91 ± 22 | 95 ± 26 | 1, 9.13 | 0.329 | 0.580 |
| ***Appetite sensations and food cravings*** | | | | | |
| Hunger, AUC | 7916 ± 2845 | 8657 ± 2397 | 1, 16.08 | 2.481 | 0.135 |
| Satiety, AUC | 8098 ± 3092 | 7638 ± 2472 | 1, 15.95 | 1.56 | 0.230 |
| PFC, AUC | 8449 ± 2380 | 9067 ± 2352 | 1, 15.84 | 1.751 | 0.205 |
| Food cravings – state | 35.1 ± 12.9 | 38.1 ± 13.4 | 1, 17.00 | 0.946 | 0.344 |
| ***Resting metabolic rate*** | | | | | |
| Resting metabolic rate, kcal/day | 1229 ± 193 | 1348 ± 155 | 1, 13.18 | 3.761 | 0.074 |

Free-living dietary intake data are the mean over 2.5 days. PFC: prospective food consumption.

**Supplementary Table 3. Descriptive data and analyses examining main and interaction effects of time and phase on measures of appetite collected in a free-living setting**

| | Late-follicular | Mid-luteal | Time | | | Phase | | | Time x Phase | | |
|---|---|---|---|---|---|---|---|---|---|---|---|
| | **Mean ± SD** | **Mean ± SD** | ***df*** | ***F*** | ***p*** | ***df*** | ***F*** | ***p*** | ***df*** | ***F*** | ***p*** |
| **Hunger, mm** | 55.2 ± 9.4 | 53.6 ± 8.4 | 2, 16.00 | 0.287 | 0.754 | 1, 16.00 | 0.145 | 0.708 | 2, 16.00 | 0.056 | 0.946 |
| **Satiety, mm** | 55.9 ± 14.3 | 57.0 ± 10.0 | 2, 16.00 | 0.906 | 0.424 | 1, 16.00 | 0.427 | 0.523 | 2, 16.00 | 1.392 | 0.277 |
| **PFC, mm** | 56.0 ± 10.6 | 56.2 ± 7.8 | 2, 16.00 | 0.280 | 0.759 | 1, 16.00 | 0.265 | 0.614 | 2, 16.00 | 0.854 | 0.444 |

Time is in increments of days (3 days) for the free-living measures of appetite. Free-living appetite ratings are expressed as mean over 3 days, with daily energy intake as a covariate in the linear mixed model.

**Supplementary Table 4. Free-living *ad libitum* dietary intake by day**

| | **Day 1** | **Day 2** | **Day 3** |
|---|---|---|---|
| ***Late-follicular*** | | | |
| Energy intake, kcal | 1069 ± 245 | 1722 ± 510 | 2194 ± 723 |
| Carbohydrate intake, g | 124 ± 37 | 208 ± 56 | 260 ± 96 |
| Fat intake, g | 46 ± 16 | 62 ± 23 | 87 ± 35 |
| Protein intake, g | 45 ± 11 | 86 ± 37 | 96 ± 27 |
| ***Mid-luteal*** | | | |
| Energy intake, kcal | 1016 ± 367 | 2085 ± 343 | 2393 ± 660 |
| Carbohydrate intake, g | 119 ± 51 | 236 ± 45 | 299 ± 70 |
| Fat intake, g | 41 ± 23 | 87 ± 20 | 90 ± 35 |
| Protein intake, g | 44 ± 14 | 96 ± 30 | 99 ± 42 |

Day 1 dietary intake only included food consumed after the study day visit.
